# Ketogenic diet reduces a neurobiological craving signature in alcohol use disorder

**DOI:** 10.1101/2023.09.25.23296094

**Authors:** Corinde E. Wiers, Peter Manza, Gene-Jack Wang, Nora D. Volkow

## Abstract

**Background and Aims:** Increasing evidence suggests that a ketogenic (high-fat, low-carbohydrate) diet intervention reduces alcohol withdrawal severity and alcohol craving in individuals with alcohol use disorder (AUD) by shifting brain energetics from glucose to ketones. We hypothesized that the ketogenic diet would reduce a brain craving signature when individuals undergoing alcohol detoxification treatment were exposed to alcohol cues.

**Methods:** We performed a secondary analysis of functional magnetic resonance data of n=33 adults with an AUD were randomized to a ketogenic diet (n=19) or a standard American diet (n=14) and underwent three weeks of inpatient alcohol detoxification treatment. Once per week, participants performed an alcohol cue-reactivity paradigm with functional magnetic resonance imaging. We extracted brain responses to food and alcohol cues and quantified the degree to which each set of brain images shared a pattern of activation with a recently validated ‘Neurobiological Craving Signature’ (NCS). We then performed a group-by-time repeated measures ANOVA to test for differences in craving signature expression between the dietary groups over the three-week treatment period. We also correlated these expression patterns with self-reported wanting ratings for alcohol cues.

**Results:** For alcohol relative to food cues, there was a main effect of group, such that the ketogenic diet group showed lower NCS expression across all three weeks of treatment. The main effect of time and the group-by-time interaction were not significant. Self-reported wanting for alcohol cues reduced with KD compared to SA but did not correlate with the NCS score.

**Conclusions:** A ketogenic diet reduces self-reported alcohol wanting, and induced lower brain craving signatures to alcohol cues during inpatient treatment for AUD.

## Introduction

Alcohol use disorder (AUD) is a chronic relapsing condition that accounts for 5% of deaths globally (Rehm and Imtiaz, 2016) and is characterized by high levels of craving and drinking despite negative consequences. Fewer than 4% of patients with AUD receive an FDA-approved medication to treat the disorder (Mark et al., 2009) and available treatments are not efficacious for all patients, and have clinical illimitations due to side effects, which can reduce adherence (Campbell et al., 2018;Kranzler and Soyka, 2018). Thus, there is a critical need to identify treatments that can reduce alcohol craving and consumption.

Improving brain energetics by means of a ketogenic diet (KD; high-fat, low-carbohydrate diet) intervention may be a novel avenue for treatment of AUD (Mahajan et al., 2021;Li et al., 2022). Acute alcohol intake shifts brain energetics from glucose to acetate, an alcohol metabolite (Volkow et al., 2013;Volkow et al., 2017). In heavy alcohol drinkers and individuals with AUD, low brain glucose and high acetate metabolism persist beyond acute intoxication (Jiang et al., 2013;Wang et al., 2013;Volkow et al., 2017), which we hypothesize contributes to alcohol withdrawal signs and symptoms, alcohol craving, and relapse due to low acetate availability (Mahajan et al., 2021). Ketone bodies (β-hydroxybutyrate [BHB], acetoacetate and acetone) structurally resemble acetate and provide an alternative to glucose as an energy source in the brain (Courchesne-Loyer et al., 2017). We recently found that a KD intervention reduced alcohol withdrawal severity and alcohol craving in individuals with an AUD undergoing inpatient detoxification, by elevating blood and brain ketone bodies (Wiers et al., 2021). Preclinical models have also demonstrated that a KD reduced both alcohol self-administration (Blanco-Gandía et al., 2021;Wiers et al., 2021) and the signs of alcohol withdrawal (Derr et al., 1981;Dencker et al., 2018;Bornebusch et al., 2021).

Koban et al. (2023) recently established a Neurobiological Craving Signature (NCS) based on brain reactivity to drug cues using functional Magnetic Resonance Imaging (fMRI). The NCS predicted drug and food craving, and distinguished drug users (alcohol, cigarettes, and cocaine) from non-users with 82% accuracy across imaging studies using machine learning techniques. The NCS is a whole-brain pattern of responses to cues, with prominent regions including ventromedial prefrontal and cingulate cortices, ventral striatum, temporal/parietal association areas, mediodorsal thalamus and cerebellum (Koban et al., 2023). Here, we performed a secondary analysis of fMRI alcohol cue reactivity data of the individuals with AUD who were randomized to receive a 3-week KD intervention versus a Control diet intervention during an inpatient alcohol detoxification program, as previously reported in Wiers et al (2021). The previous study reported both clinical outcomes and dorsal anterior cingulate cortical activation outcomes for alcohol > neutral cues, and food > neutral cues (Wiers et al., 2021), but not for alcohol > food contrasts. Here we computed the novel NCS expression levels for the alcohol > food cues, as per Koban et al (2023). We hypothesized lower NCS with KD compared to the Control diet, which would associate with low alcohol wanting ratings and elevated blood BHB levels.

## Methods

### Participants and Screening

This report involves a secondary analysis of the fMRI alcohol cue reactivity paradigm of a dataset previously reported (Wiers et al., 2021). Data on patient demographics, blood ketone levels, and behavioral craving ratings have been reported previously and were shown again here for context. Details of the clinical trial methodology can be found in (Wiers et al., 2021). Briefly, individuals with AUD were admitted to the National Institute on Alcohol Abuse and Alcoholism (NIAAA) inpatient unit while receiving treatment as usual, and within two days were randomized to receive either a ketogenic diet (n=19 randomized within 0.95±0.23SD days) or a standard American Control diet (n=14 randomized within 0.78±0.42SD days) for three weeks. There were no significant differences in demographic or clinical characteristics between groups (**Table 1**). Patients provided written informed consent to participate in the study, which was approved by the Institutional Review Board at the National Institutes of Health (Combined Neurosciences White Panel). Participants were MRI scanned between October 2017 and February 2020. The study was registered at ClinicalTrials.gov (NCT03255031).

**Table 1.** Demographics and Clinical Characteristics of the ketogenic diet (KD) and standard American (SA) groups.

|  | KD (*n*=19) | SA (*n*=14) | p-value |
| --- | --- | --- | --- |
| Age (years) | 39.3 (11.2) | 44.2 (16.4) | 0.311 |
| Sex | 7 females (37%)<br>12 males (63%) | 3 females (21%)<br>11 males (79%) | 0.341 |
| BMI admission | 24.5 (3.4) | 27.6 (5.4) | 0.051 |
| Study weight loss (kg) | 1.4 (2.8) | 1.8 (2.2) | 0.692 |
| Mean calories per day during study (kcal) | 2674.3 (469.2) | 2632.1 (261.5) | 0.764 |
| Ethnicity | 6 Black/African American<br>11 White, 2 Multiracial | 6 Black/African American<br>6 White, 2 Multiracial | 0.694 |
| Years of education | 12.7 (2.8) | 14.2 (2.5) | 0.110 |
| WASI IQ | 99.2 (16.3) | 98.3 (22.5) | 0.892 |
| Smoking status | 10 smokers<br>9 non-smokers | 9 smokers<br>5 non-smokers | 0.503 |
| LDH (kg) | 984.9 (945.5) | 1414.4 (1462.6) | 0.320 |
| TLFB Drinks/day | 15.2 (8.4) | 17.0 (9.8) | 0.584 |
| ADS | 21.0 (9.8) | 23.6 (7.2) | 0.299 |

Abbreviations: ADS, Alcohol Dependence Scale, BMI, Body Mass Index, IQ, Intelligence Quotient, LDH, Lifetime Drinking History, TLFB, Timeline Followback, WASI, Wechsler Abbreviated Scale of Intelligence.

Patients were screened to exclude individuals who had contraindications for MRI, major medical problems, chronic use of psychoactive medications, liver disease, head trauma, or milk or soy allergy; and current diagnosis of a substance use disorder (SUD) (other than AUD or nicotine) that was severe or any other major psychiatric disorder that needed treatment for longer than a month in the past year as assessed by the Structured Clinical Interview for the Diagnostic and Statistical Manual of Mental Disorders (DSM-IV or V) (American Psychiatric Association, 2013). Participants had at least five years’ history of heavy drinking (5+ drinks/day or 4+ drinks/day on at least five days per month for men or women, respectively). Alcohol had to be specified as the preferred drug of choice. All participants were free of psychoactive medications within 24 hours of study procedures (except benzodiazepines as needed for detoxification) and had a negative urine drug screen (and, if needed, a negative saliva screen to rule out THC-COOH use, Draeger DrugTest5000; Draeger Safety Diagnostics, Lübeck, Germany) on days of testing.

### Diet intervention

The KD and Control diet meals were equicaloric. The KD provided a classic 4:1 ratio of grams of fat: grams of carbohydrate and protein (i.e., 80% fat, 15 % protein, 5% carbohydrates). The standard American Control diet corresponded to 50% calories from carbohydrates, 15% protein and 35% fat. For each meal at breakfast, lunch and dinner, the diets consisted of a shake (either KD or Control diet) that provided 75% of meal calories, and a ketogenic solid snack (e.g., scrambled egg, yogurt with nuts, salad, chicken salad, broth) that provided 25% of meal calories, which ensured double blinding (Wiers et al., 2021). Patients were allowed to drink water *ad libitum* and were given the option to drink tea or coffee with or without stevia, and/or diet ginger-ale with their meals. Meals were provided from the Nutrition Department Metabolic Kitchen with all foods and beverages weighed on a gram scale. There was no effect of group on diet expectation, indicating that the blinding of the diet was successful (Wiers et al., 2021).

### Blood ketone measures

BHB ketone levels were assessed on the day of informed consent (baseline), and before breakfast on the morning of each MRI scan in week 1, 2, and 3, using a finger stick and Precision Xtra Blood Ketone Monitoring System strips (Abbott; Alameda, CA).

### Questionnaires and ratings

At screening, participants completed the Wechsler Abbreviated Scale of Intelligence (WASI-II) subtests Matrix Reasoning and Vocabulary as a proxy for general intelligence (Wechsler, 1999), the Timeline Followback (TLFB) to assess daily alcohol consumption in the 90 days prior to the study (Sobell and Sobell, 1996), the Lifetime Drinking History (LDH) to assess lifetime alcohol consumption (Skinner and Sheu, 1982), the Alcohol Dependence Scale (ADS) to assess severity of dependence (Skinner and Allen, 1982).

On a weekly basis, participants rated their alcohol craving on the Desire for Alcohol Questionnaire (DAQ) (Love et al., 1998).

### MRI acquisition, and Preprocessing

Participants underwent three MRI scans, in the first, second and third week of diet initiation. MRIs were performed on a 3.0T Magnetom Prisma scanner (Siemens Medical Solutions USA, Inc., Malvern, PA) equipped with a 32-channel head coil. T1-weighted 3D magnetization-prepared gradient-echo (MP-RAGE, TR/TE=2200/4.25 ms; FA=9°, 1 mm isotropic resolution) and T2-weighted multi-slice spin-echo (TR/TE=8000/72ms; 1.1 mm in-plane resolution; 94 slices, 1.7-mm slice thickness; matrix=192) pulse sequences were used to acquire high-resolution anatomical brain images. One participant did not complete session 3 due to scheduling problems. For functional MRI, a 32-channel head coil and a standard echo planar imaging (EPI) sequence were used: sequential interleaved acquisition, repetition time 1.5s, echo time 30 ms, flip angle α=70°, 64×64 pixels in-plane resolution, 36 slices, slice thickness 4 mm, voxel dimensions 3×3×4 mm^3^, field of view 192×192 mm^2^. Stimuli were presented on a black background under dimmed room lighting using a liquid-crystal display screen (BOLDscreen 32, Cambridge Research Systems; UK).

#### fMRI cue reactivity

A total of 40 alcohol, 40 appetitive food, 40 neutral images were randomly presented in an event-related design using E-prime software (see Wiers et al. (2020), for details). There were 3 runs showing 40 cues that included all 3 categories of cues (alcohol, food, neutral). Cues were presented at 750ms. Pictures were selected from the International Affective Picture System (IAPS) and from in-house pictures libraries. To ensure that participants were paying attention they were instructed to press a button if they saw a bicycle. Total task duration was 13 min and 30 sec. After completion of the MRI sessions, participants were asked to rate the cues for their subjective valence (“How negative/positive do you find the picture?”, -3 to 3 [very negative – very positive]) and wanting (“How much do you want to consume this right now?” from 0 to 6 [not at all – extremely]), on a 7-point Likert scale.

#### Preprocessing

Functional data analysis was performed with SPM8 (Welcome Department of Cognitive Neurology, London, UK). During preprocessing, scans were slice-time corrected, spatially realigned, co-registered to the T1 structural images and normalized to the Montreal Neurological Institute (MNI) template. Smoothing was performed with a 6mm full-width at half-maximum Gaussian kernel.

### Neurobiological Craving Score

For fMRI, 3 fMRI regressors – food, alcohol, and neutral events were created and then convolved with the hemodynamic response function with default temporal filtering of 128 seconds. Six realignment parameters were included as regressors of no interest. The following contrasts were calculated at the single-subject level: alcohol>baseline (average), food>baseline, for which dorsal anterior cingulate activation has been reported previously for KD versus the Control diet (Wiers et al., 2021). In a secondary analysis, we took the first-level whole brain maps for each individual, and performed dot-product multiplication between the NCS map from Koban et al. (2023) and the contrast alcohol > food cues. If the resulting NCS ‘expression level’ is high, it indicates a brain response pattern to alcohol > food cues that is similar to the NCS map which predicted alcohol and drug craving.

## Statistical Analysis

For all measures, we performed mixed ANOVAs with group (KD/Control diet) as between-group factor and time as within-subjects factor using SPSS. Post hoc t-tests were performed with significance threshold of α<0.05.

## Results

### Serum Beta-hydroxybutyrate levels

The 4:1 KD intervention significantly increased serum BHB levels compared to the Control Diet (group × time interaction: F_3,93_=67.0, p<0.0001, *η*^*2*^=0.68). Blood ketones did not differ between groups at baseline before diet initiation (t_31_=1.4, p=0.2), and became significantly elevated in the KD group the morning of the MRI day in week 1 (mean BHB in KD group=1.6±1.5 mM), week 2 (mean KD=4.6±1.0mM), and week 3 (mean KD=4.4±1.5mM) (all p<0.001; **Figure 1**), indicative of nutritional ketosis. The slight decrease in week 3 compared to week 2 for blood measures reflected two participants who were not compliant with the KD in week 3, one for 1 day and the other for 2 days.

**Figure 1.**
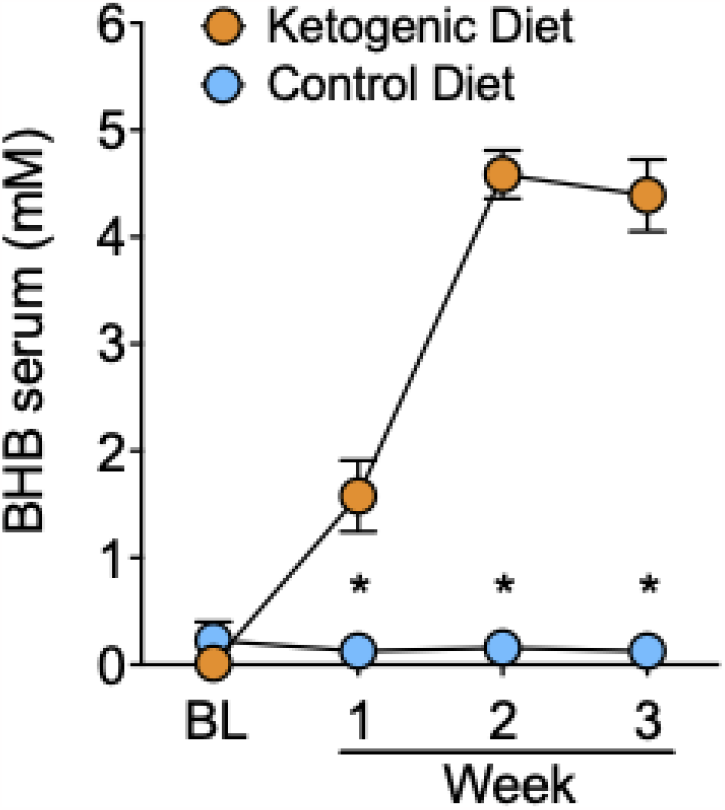
Serum levels of BHB in the Ketogenic and Control Diet group. \**p* < 0.001, KD different from Control diet.

All participants on the Control diet demonstrated blood BHB levels of <0.2mM, indicating compliance to the Control diet, and no presence of nutritional ketosis.

### Self-reported Alcohol Craving

Self-reported DAQ alcohol craving showed a trend for a main effect of time (F_2,58_=2.5, p=0.09, *η*^*2*^=0.08), which was largely due to reductions in DAQ scores in the KD group (F_2,32_=3.2, p=0.056, *η*^*2*^=0.17), but not the Control diet group (F_2,26_=0.45, p=.64, *η*^*2*^=0.03). The effect of group or group × time for DAQ did not reach statistical significance.

### Alcohol cue reactivity

Mean ratings of “wanting” alcohol cues relative to the neutral cues used in the fMRI cue reactivity task, decreased over time in the KD (F_2,32_=6.6, p=0.004, *η*^*2*^=0.29) but not in the Control diet group (F_2,24_=0.5, p=0.62, *η*^*2*^=0.04), and the group × time interaction effect was significant (F_2,56_=4.9, p=0.048, *η*^*2*^=0.20; **Figure 2A**). There were no significant group effects for “wanting” ratings when exposed to food cues *versus* neutral cues (**Figure 2A**), and no effects for valence ratings of alcohol cues or food cues, relative to neutral cues.

**Figure 2.**
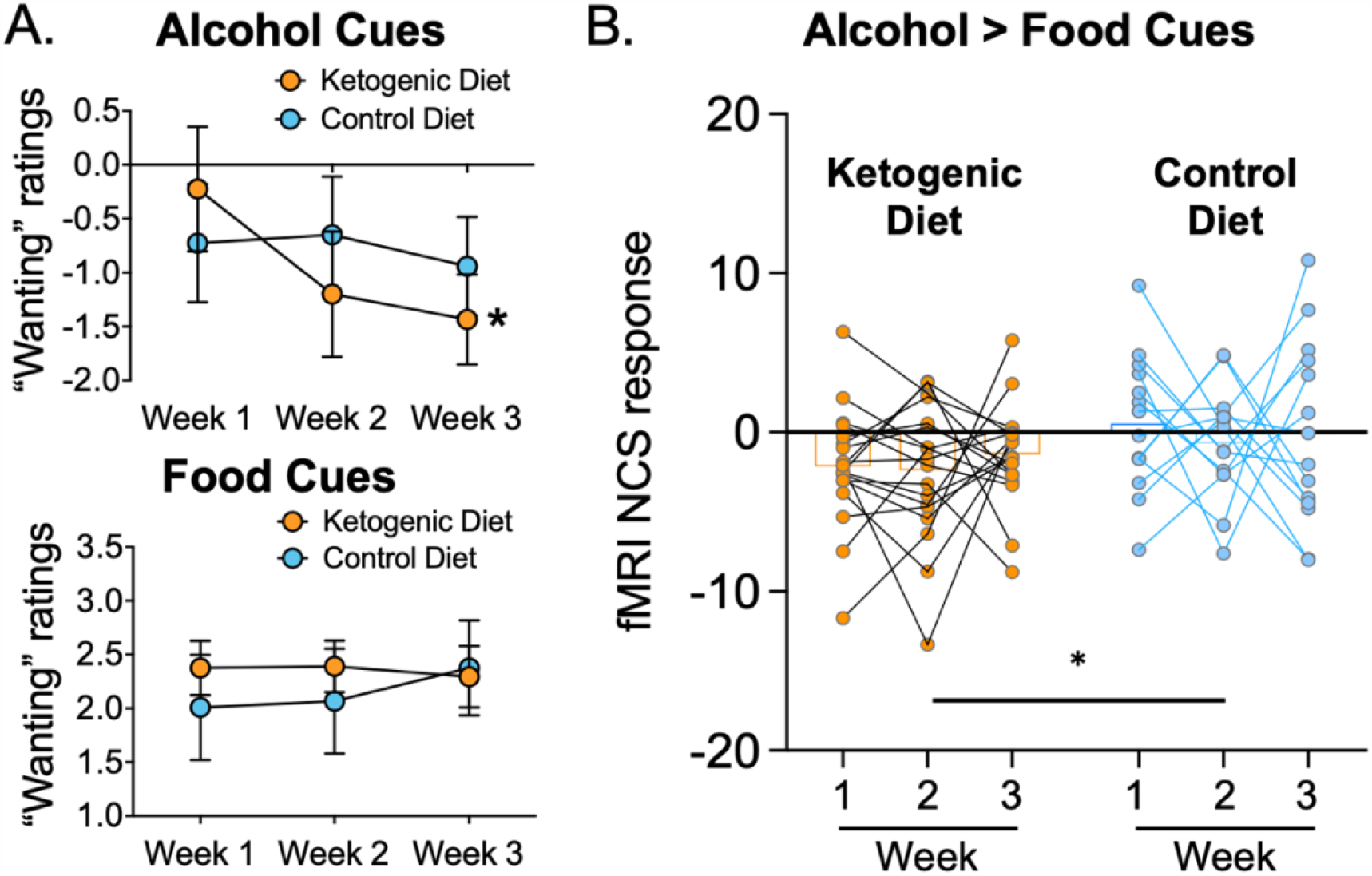
**A)** Alcohol “wanting” ratings of alcohol cues were lower in KD than Control diet after diet initiation. There were no group differences for “wanting” ratings caused by food-cues. **B)** The fMRI NCS response was lower in the KD group than the Control Diet throughout the 3 weeks of dietary intervention.

### Neurobiological Craving Score

There was a main effect of group on the alcohol > food NCS (F_1,30_=9.9, p=0.004, *η*^*2*^=0.248), with lower overall NCS in the KD group than the Control diet (**Figure 2B**). The effect of group × time or main effect of time for NCS did not reach statistical significance. Average 3-week NCS scores correlated with average 3-week serum BHB levels (r=-0.365, p=0.037), corroborating that higher ketone levels covaried with lower neurobiological craving for alcohol in AUD patients. There were no significant associations between the alcohol NCS with alcohol craving or wanting.

## Discussion

We applied a newly identified fMRI Neurobiological Craving Signature (NCS) for drug craving (Koban et al., 2023) on a previously reported fMRI alcohol cue reactivity dataset acquired from individuals with AUD who were randomized into a 3-week ketogenic diet (KD) or control diet interventions during inpatient alcohol detoxification (Wiers et al., 2021). The KD group demonstrated lower NCS responses compared to the Control diet throughout the 3-week intervention, which covaried with the average blood BHB levels on the days of the MRI scans. Moreover, the KD compared to Control diet group demonstrated lower “wanting” ratings of the alcohol cues presented in the alcohol cue reactivity task, which did not correlate with NCS. These findings provide novel insight into the effects of KD on neural signatures of alcohol craving, and add to the previously reported effects of KD increasing dorsal cingulate cortical activation for both alcohol and food cues. Overall, these findings provide evidence for the potential therapeutic benefit of a KD in reducing alcohol wanting and neurobiological craving for alcohol.

These findings are in line with recent preclinical models of alcohol dependence that showed that a KD reduced alcohol self-administration (Blanco-Gandía et al., 2021;Wiers et al., 2021) and the signs of alcohol withdrawal (Derr et al., 1981;Dencker et al., 2018;Bornebusch et al., 2021). The reductions in alcohol NCS in the KD group may be the consequence of the reduced withdrawal symptoms and need for benzodiazepines in the KD group as reported previously (Wiers et al., 2021), or may reflect a potentially beneficial effect of KD in disrupting conditioning to alcohol through its provision of ketone bodies that can serve as energy substrates. Although most of the research studying conditioned effects of alcohol associate it to its reinforcing effects mediated by its enhancement of GABAergic, opioid and dopaminergic systems, it is possible that the KD-induced bioenergetic switch from glucose to ketones is also relevant to its reinforcing effects, particularly given that energy-rich stimuli engender strong conditioned responses (de Araujo et al., 2008;Rejeski et al., 2010;Volkow et al., 2017). In line with this, the NCS scores were negatively correlated with serum BHB levels of participants. We thus speculate that a KD would satisfy the altered energy requirements associated with chronic alcohol consumption through its metabolism of BHB. Alternatively, ketone supplementation decreases ghrelin levels and appetite levels (Stubbs et al., 2018), which in turn may have reduced alcohol craving and brain reactivity to alcohol cues (Farokhnia et al., 2017).

Our findings confirm our overall hypothesis that providing ketone bodies via a KD intervention during detoxification would attenuate the emergence of alcohol craving. We reasoned that the abrupt transition from the brain’s consumption of ketone bodies, which occurs in AUD as an adaptation to repeated alcohol intake, to the use of glucose as energy source, which reemerges with detoxification, may contribute to the alcohol withdrawal syndrome (Volkow et al., 2013). Notably, BHB, which is a major ketone body is thought to be a more efficient ‘fuel’ source than glucose. Therefore, inducing ketosis with a KD during detoxification would bolster or halt such an abrupt transition (Courchesne-Loyer et al., 2017), thereby reducing alcohol withdrawal signs and symptoms, alcohol craving, and conditioned brain reactivity to alcohol cues. Disorders of brain glucose metabolism are associated with disrupted neuronal excitability, including epilepsy (Pascual et al., 2008), and are ameliorated by KD. Thus, the reduction of withdrawal symptoms by KD may reflect an attenuation of neuronal excitability following alcohol discontinuation (Long et al., 2017).

This study is, to our knowledge, the first to examine the effect of inpatient substance use disorder treatment on the NCS. Establishing a biomarker of craving has been a highly sought after target in neuroscience and psychiatry (Kronberg and Goldstein, 2023). Addiction is a chronic condition, and even in recovery, an individual will experience strong fluctuations in vulnerability to relapse (Yip and Konova, 2022). However, abstinence and drug use themselves are often not the best outcome measures to predict subsequent relapse; therefore, a neuroimaging biomarker that could better track recovery would be a highly useful tool for identifying when to intervene and prevent future drug use (Goldstein, 2022). The NCS, along with other recent machine learning-based neural signatures of craving (Garrison et al., 2023) represent an exciting new approach for tracking risk of relapse during recovery. The current findings suggest that the NCS may indeed be sensitive to substance use disorder intervention and the recovery process, complementing recent studies using decision-making behavior to predict craving fluctuations in an opioid use disorder sample (Biernacki et al., 2022). Future markers that combine imaging and behavioral markers may improve prediction further.

The main limitation of our study is that the first MRI was performed 4.0±1.7SD days after inpatient admission (no group differences) at a time when the KD participants were already in mild ketosis (blood BHB=1.6±1.5 mM). It was not possible to perform MRI scans at an earlier time point before diet initiation, due to clinical constraints of participants being in acute alcohol withdrawal. Our design therefore did not have an absolute MRI baseline that allowed for comparison of NCS before and after diet initiations. This may be why our study found a main effect of KD on the alcohol NCS, but no group x diet interaction effect, as expected. Future studies are needed that test the effects of ketone supplementation in participants with AUD that would include a baseline measure. Another limitation of our clinical study was the relatively small sample size, which was constrained by the complexity of the study that required 3 weeks of hospitalization and compliance with the diet protocol. Nevertheless, we demonstrated the first clinical trial on a ketogenic diet intervention in inpatients with AUD undergoing detoxification, with a complex and successful randomized and blinded design.

In sum, this study documents an effect of KD in reducing a neurobiological signature associated with alcohol craving in individuals with AUD. These findings are in line with recent preclinical data that show that a KD (Blanco-Gandía et al., 2021) and a history of KD (Wiers et al., 2021) reduce acute self-administration in alcohol-dependent rodents. It remains to be studied whether a KD decreases alcohol consumption in a human population with AUD.

## Data Availability

All data produced in the present study are available upon reasonable request to the authors

## Acknowledgments

We thank Nancy Diazgranados, Leandro Vendruscolo, Karen Torres, Minoo McFarland, Lori Talagala, Michele-Vera Yonga, Yvonne Horneffer, David George, Tonette Vinson, Ehsan Shokri-Kojori, Dardo Tomasi, Rui Zhang, Sukru Baris Demiral, Mackenzie Cervenka, Sara Turner, Shanna Yang, Todd King and Richard Veech for their contributions and discussions. All data needed to evaluate the conclusions in the paper are present in the paper and/or the Supplementary Materials.

## Funding

This work was accomplished with support from the National Institute on Alcohol Abuse and Alcoholism, Intramural Research Program (Y1AA-3009 to Dr. Volkow). Dr. Wiers was supported by a NARSAD Young Investigator Grant (28778), and a K99/R00 grant AA026892.

## Competing interests

The authors declare no conflicts of interest.

## Notes

### Competing Interest Statement

The authors have declared no competing interest.

### Clinical Trial

NCT03255031

### Author Declarations

Patients provided written informed consent to participate in the study, which was approved by the Institutional Review Board at the National Institutes of Health (Combined Neurosciences White Panel). Participants were MRI scanned between October 2017 and February 2020. The study was registered at ClinicalTrials.gov (NCT03255031).

